# Application of BRACE Method to Address Treatment Selection Bias in Observational Data

**DOI:** 10.1101/2021.05.17.21257332

**Authors:** Casey W. Williamson, Tyler Nelson, Caroline A. Thompson, Lucas K. Vitzthum, Kaveh Zakeri, Paul Riviere, Alex K. Bryant, Andrew B. Sharabi, Jingjing Zou, Loren K. Mell

## Abstract

**Background:** Cancer treatments can paradoxically appear to reduce the risk of non-cancer mortality in observational studies, due to residual confounding from treatment selection bias. Here we apply a novel method, Bias Reduction through Analysis of Competing Events (BRACE), to reduce bias in the presence of residual confounding.

**Methods:** We studied 36630 prostate cancer patients, 4069 lung cancer patients, and 7117 head/neck cancer patients, using the Veterans Affairs Informatics and Computing Infrastructure database. We estimated effects of intensive treatment (prostate: prostatectomy vs. radiotherapy; lung: lobectomy vs. sublobar resection or radiotherapy; head/neck: radiotherapy with concurrent cisplatin and/or multiagent induction vs. radiotherapy with or without alternative systemic therapy) on cancer-specific mortality, non-cancer mortality, and overall survival (OS), using both multivariable Cox (MVA) and propensity score (inverse probability treatment weighting (IPTW)) models. Next, we applied the BRACE method to adjust for residual confounding, based on the observed treatment effect on competing event and relative event hazards.

**Results:** For each cohort, intensive treatment was associated with significantly reduced hazards for cancer-specific mortality, non-cancer mortality, and OS. Compared to the results for MVA and IPTW models, hazard ratios (95% confidence intervals) for the effect of intensive treatment on OS were attenuated in each cohort after applying BRACE: (prostate-MVA: 0.75 (0.71, 0.80), IPTW: 0.73 (0.66, 0.75), BRACE: 0.98 (0.95, 1.00); lung-0.79 (0.68, 0.91), 0.79 (0.66, 0.89), BRACE: 0.81 (0.65, 0.94); head/neck-0.71 (0.66, 0.76), 0.70 (0.66, 0.76), BRACE: 0.81 (0.76, 0.86)). BRACE estimates were similar to findings from meta-analyses and randomized trials.

**Conclusions:** We found evidence of residual confounding in several observational cohorts after applying standard methods, which were mitigated after applying BRACE. Application of this method could provide more reliable estimates and inferences when residual confounding is identified and represents a novel approach to improving the validity of outcomes research.

## INTRODUCTION

Bias due to residual confounding (often called treatment selection bias) is an important issue when drawing inferences from non-randomized comparative effectiveness studies.^1,2^ In observational data, multivariable regression models and propensity scores are common approaches to reduce bias from measured confounders.^3^ However, residual confounding from unmeasured or unknown confounders remains a pernicious problem that can undermine conclusions from such analyses and cannot be overcome by adjustment, scoring, or weighting methods.^4-8^ Importantly, biased inferences from observational data can mislead the medical field, resulting in patients receiving toxic, costly, and ineffective therapies.^9,10^

Competing event analysis allows for identification of residual confounding problems in observational data, particularly when the effect of a treatment on competing events can be bounded *a priori*.^11^ For example, while the addition of a novel cancer treatment to a standard regimen may have no effect on or even increase mortality from non-cancer health events, such as cardiac disease, it should not intrinsically *reduce* the incidence of such non-cancer events. Despite this, in non-randomized data, competing event analysis can reveal a *lower* incidence of competing health events in the group receiving intensified treatment, due to unmeasured confounding by more favorable health characteristics in this group, even after appropriately controlling for measurable confounders.^12^ When present, this phenomenon typically indicates the presence of residual confounding, assuming that more intensive treatment does not truly reduce the risk for competing events.

While diagnosing residual confounding with a competing events analysis is helpful,^12^ there remains no consensus on how to address it.^2,4^ Here we apply a novel method, Bias Reduction through Analysis of Competing Events (BRACE), to mitigate this bias. We previously showed that BRACE reduces bias and model error in simulated data.^13^ Here we sought to test the performance of this method in three large observational cohorts of U.S. veterans treated for prostate, lung, or head/neck cancer and to compare our findings to results from landmark randomized trials and/or meta-analyses. We also applied BRACE to externally collected data for further validation.

## METHODS

### Population and Sampling Methods

We applied BRACE to observational cohorts of patients treated for prostate cancer, lung cancer, and head/neck cancer, sampled from the Veterans Affairs (VA) Informatics and Computing Infrastructure (VINCI) database. VINCI contains detailed electronic medical records for veterans treated across the United States with tumor registry data collected by trained registrars according to standardized protocols.^14^ Further details on each cohort are provided below. This study was approved by our institutional and local VA institutional review boards. Waiver of informed consent was obtained.

### Outcomes

The primary outcome of interest was overall survival (OS). For competing risks analysis, we analyzed two events: cancer-specific mortality and non-cancer (competing) mortality. Patients with documented follow-up visits and no death event were coded as alive at last follow-up with event times censored. Date of death and cause of death were obtained via the National Death Index from the Department of Defense for deaths through 2014 and tumor registry data for deaths after 2014, which are linked to the VA data by social security numbers. Survival times (days) were measured from the date of diagnosis.

### Prostate Cohort

The prostate cancer cohort included 36,630 patients with cT1-T2 cancer of the prostate, with prostate-specific antigen (PSA) < 20, who were diagnosed between 2000 and 2015 and received radical prostatectomy and/or RT. The primary treatment effect of interest was radical prostatectomy vs definitive radiotherapy. Covariates were age (categorical in 10-year increments), race (White/Black/Other), Charlson comorbidity index (CCI, 0/1/≥2), body mass index (BMI, <18.5, 18.5-25, 25-30, >30), smoking status (nonsmoker/current smoker), employment status (not employed/employed), marital status (not married/married), alcohol use (nondrinker vs current use), baseline PSA (<10 vs. 10-20), Gleason Score (6/7/8-10), and T category. Each model was stratified by use of hormonal therapy. Use of hormonal therapy was recorded a binary yes/no based on whether hormonal therapy was given within 6 months of RT/surgery; duration of hormonal therapy could not be ascertained. Alcohol use was excluded from each model after backward selection.

For additional external validation (i.e., in a cohort where we did not determine the sampling methods), we applied the BRACE method to a cohort of patients from SEER data with low-risk prostate cancer (cT1-T2a, PSA <10, and Gleason 6) who received radical prostatectomy, brachytherapy, or external beam radiotherapy from 2005-2015, as described in detail elsewhere.^15^ Brachytherapy and external beam radiotherapy were combined in a single radiation therapy group and were compared to radical prostatectomy. For multivariable analysis, initial variables included age at diagnosis (continuous), race (White/Black/Other/Unknown), insurance status (Insured/Uninsured/Unknown), and year of diagnosis (continuous). Year of diagnosis was dropped on backward model selection (p>0.2). On balance check after propensity score weighting, all covariates had a standardized mean difference of <0.05 except for age (0.11).

### Lung Cohort

The lung cohort included 4,069 patients with biopsy-proven clinical stage I (T1 or T2a, N0) non-small cell lung cancer (NSCLC) diagnosed between 2006 and 2015 and treated definitively with surgery (lobectomy or sublobar resection) or RT, as previously described.^16^ The primary treatment effect of interest for this cohort was lobectomy vs. sublobar resection or definitive RT. Missing variables were imputed using iterative robust model-based imputation (IRMI)^17^. Covariates were age (categorical in 10-year increments), sex, race (White/Black/Other), smoking status (never/current/past), CCI (0/1/2/3+), pretreatment forced expiratory volume in one second (FEV1) (pre-treatment percent predicted, categorical, <30%, 31-50%, 51-80%, >80%), T category (T1a vs T1b vs T2a), grade, and histology. Smoking status and histology were excluded from the models for OS and CSM after backward selection. Grade was excluded from the NCM model after backward selection.

### Head/Neck Cohort

The head and neck cohort included 7,117 patients with locoregionally advanced, non-metastatic (AJCC 7th edition stage III-IVB) squamous cell carcinoma of the oropharynx, oral cavity, larynx, and hypopharynx diagnosed between 2005 and 2015 and treated with definitive (at least 5 weeks of) radiation therapy (RT) with or without chemotherapy, as previously described.^18^ The primary treatment effect of interest for this cohort was intensive therapy (defined as RT with concurrent cisplatin or with multiagent induction chemotherapy) vs. alternative therapy (defined as RT alone or RT with other systemic therapies not qualifying as intensive), as previously described. Covariates in each model were age (categorical), sex, race (White/Black/Other), Eastern Cooperative Oncology Group (ECOG) performance status (2 vs. 0-1), CCI (2 vs. 0-1), BMI (<18.5, 18.5-25, 25-30, >30), marital status (currently employed vs. not employed), employment status (currently employed vs. not employed), primary tumor site (oral cavity vs. hypopharynx/larynx vs. p16-negative oropharynx vs. p16-positive oropharynx), T category (1-2 vs 3-4), and N category (N0-1 vs 2-3) per American Joint Committee on Cancer (AJCC) 7th edition. Unknown p16 status was considered positive for oropharyngeal cancer patients and negative for non-oropharyngeal cancer patients, consistent with other studies.^19,20^ Other missing variables were imputed using IRMI. For the CSM model, sex was excluded after backward selection. For the NCM model, race and employment status were excluded after backward selection.

### Statistical Analysis

Unadjusted and multivariable Cox proportional hazards models (MVA) were fit for each outcome and each cohort. Adjustment variables were determined using backward selection, retaining covariates found to be associated with OS (threshold: p < 0.20). The proportional hazards assumption was checked (*cox*.*zph* function in the *survival* package in R), and when violated, treatment was modeled as a time-varying covariate. For propensity score adjustment, we implemented inverse probability treatment weighting (IPTW) with multivariable Cox models using stabilized weights derived from the same sets of covariates. An average treatment effect (ATE) approach was used for estimation of treatment effects. Each IPTW model was checked for covariate balance across treatment groups, with a standardized mean difference (SMD) threshold of 0.05. Statistical analyses were performed using R version 4.0.2.

### BRACE Technique

BRACE-corrected estimates of the effect of treatment on overall survival (OS,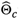 were obtained as the sum of the (adjusted) effect estimate on OS (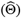) and the product 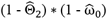,where 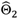 is the adjusted effect estimate on the competing event (NCM) and 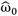 is the estimated proportion of the overall event hazard attributable to the primary event (CSM),^11-12, 22-24^ i.e.:

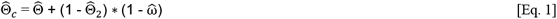

BRACE was then applied to the IPTW model estimates to obtain bias-corrected estimates 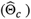. Bootstrapped confidence intervals for 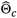 were estimated with 500 replicates. Monte Carlo estimates of 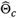 were obtained by randomly co-sampling values of 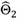 and 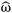 from their respective distributions (1000 replicates). Confidence intervals were defined by the 2.5^th^ and 97.5^th^ percentiles of the sampling distributions. The BRACE method does not generate p-values; confidence intervals were compared between methods. More details regarding BRACE derivation were described previously.^13^

## RESULTS

There were 36630 patients in the prostate cohort (**Table 1**), 4069 patients in the lung cohort (**Table 2**), and 7117 patients in the head and neck cohort (**Table 3**). On balance check for each IPTW model, all covariables had a mean difference of < 0.05, indicating appropriate balance after weighting by propensity score (**Supplemental Figure 1**). Results for standard approaches using either multivariable Cox models or IPTW models were largely similar and are presented in tabular form, with comparison of IPTW vs. BRACE-corrected estimates emphasized in the text.

**Table 1.** Descriptive statistics for prostate cancer cohort. All variables listed in format of number of patients (column percentage) except as otherwise noted. PSA = prostate-specific antigen, SD= standard deviation.

| COVARIATES | RADIATION<br>(N= 21615) | PROSTATECTOMY<br>(N= 15015) | OVERALL<br>(N= 36630) |
| --- | --- | --- | --- |
| **AGE** |  |  |  |
| <50 | 177 (0.8) | 442 (2.9) | 619 (1.7) |
| 50-59 | 3597 (16.6) | 4393 (29.3) | 7990 (21.8) |
| 60-69 | 11033 (51.0) | 8248 (54.9) | 19381 (52.6) |
| 70-79 | 6364 (29.4) | 1679 (11.2) | 8043 (22.0) |
| ≥80 | 444 (2.1) | 253 (1.7) | 697 (1.9) |
| **RACE** |  |  |  |
| WHITE | 13952 (64.5) | 10173 (67.8) | 24125 (65.9) |
| BLACK | 6505 (30.1) | 3948 (26.3) | 10453 (28.5) |
| OTHER | 1158 (5.4) | 894 (6.0) | 2052 (5.6) |
| **CHARLSON COMORBIDITY INDEX** |  |  |  |
| 0 | 15008 (69.4) | 11837 (78.8) | 26845 (73.3) |
| 1 | 3798 (17.6) | 1903 (12.7) | 5701 (15.6) |
| ≥2 | 2809 (13.0) | 1275 (8.5) | 4084 (11.1) |
| **BODY MASS INDEX** |  |  |  |
| UNDERWEIGHT (<18.5) | 542 (2.5) | 297 (2.0) | 839 (2.3) |
| NORMAL (≥18.5, <25) | 5022 (23.2) | 3373 (22.5) | 8395 (22.9) |
| OVERWEIGHT (≥25, <30) | 7594 (35.1) | 5971 (39.8) | 13565 (37.0) |
| OBESE (≥30) | 8457 (39.1) | 5374 (35.8) | 13831 (37.8) |
| **CURRENT SMOKER** |  |  |  |
| YES | 14286 (66.1) | 9444 (62.9) | 23730 (64.8) |
| NO | 7329 (33.9) | 5571 (37.1) | 12900 (35.2) |
| **CURRENTLY EMPLOYED** |  |  |  |
| YES | 3595 (16.6) | 3784 (25.2) | 7379 (20.1) |
| NO | 18020 (83.4) | 11231 (74.8) | 29251 (79.9) |
| **CURRENTLY MARRIED** |  |  |  |
| YES | 11157 (51.6) | 8525 (56.8) | 19682 (53.7) |
| NO | 10458 (48.4) | 6490 (43.2) | 16948 (46.3) |
| **STAGING PSA (NG/ML, MEAN, SD)** | 6.94 (3.67) | 6.15 (3.35) | 6.62 (3.56) |
| **CLINICAL T CATEGORY** |  |  |  |
| 1 | 15424 (71.4) | 10815 (72.0) | 26239 (71.6) |
| 2A | 3067 (14.2) | 2241 (14.9) | 5308 (14.5) |
| 2B | 1093 (5.1) | 539 (3.6) | 1632 (4.5) |
| 2C | 2031 (9.4) | 1420 (9.5) | 3451 (9.4) |
| **STAGING GLEASON SCORE** |  |  |  |
| 6 | 8137 (37.6) | 5148 (34.3) | 13285 (36.3) |
| 7 | 9734 (45.0) | 7618 (50.7) | 17352 (47.4) |
| 8 | 2386 (11.0) | 1355 (9.0) | 3741 (10.2) |
| 9-10 | 1358 (6.3) | 894 (6.0) | 2252 (6.1) |
| **RISK GROUP STRATIFICATION** |  |  |  |
| LOW | 6476 (30.0) | 4309 (28.7) | 10785 (29.4) |
| INTERMEDIATE | 11395 (52.7) | 8457 (56.3) | 19852 (54.2) |
| HIGH | 3744 (17.3) | 2249 (15.0) | 5993 (16.4) |
| **RECEIPT OF HORMONE THERAPY** |  |  |  |
| YES | 6592 (30.5) | 674 (4.5) | 7266 (19.8) |
| NO | 15023 (69.5) | 14341 (95.5) | 29364 (80.2) |

**Table 2.** Descriptive statistics for lung cancer cohort. All variables listed in format of number of patients (column percentage). SABR= Stereotactic Ablative Radiotherapy, FEV1= Forced Expiratory Volume in 1 second, percent predicted.

| LUNG VARIABLES | SABR<br>(N= 449) | SUBLOBAR RESECTION<br>(N= 634) | LOBECTOMY<br>(N= 2986) | OVERALL<br>(N= 4069) |
| --- | --- | --- | --- | --- |
| **AGE** |  |  |  |  |
| <50 | < 10 | < 10 | 43 (1.4) | 49 (1.2) |
| 50-59 | 25 (5.6) | 81 (13.1) | 533 (17.8) | 641 (15.8) |
| 60-69 | 181 (40.3) | 268 (42.3) | 1533 (51.3) | 1982 (48.7) |
| 70-79 | 161 (35.9) | 207 (32.6) | 722 (24.2) | 1090 (26.8) |
| ≥80 | 81 (18.0) | 71 (11.2) | 155 (5.2) | 307 (7.5) |
| **RACE** |  |  |  |  |
| WHITE | 379 (84.4) | 535 (84.4) | 2519 (84.4) | 3433 (84.4) |
| BLACK | 63 (14) | 85 (13.4) | 404 (13.5) | 552 (13.6) |
| OTHER | < 10 | 14 (2.2) | 63 (2.1) | 84 (2.1) |
| **SEX** |  |  |  |  |
| FEMALE | 12 (2.7) | 26 (4.1) | 126 (4.2) | 164 (4.0) |
| MALE | 437 (97.3) | 608 (95.9) | 2860 (95.8) | 3905 (96.0) |
| **CHARLSON COMORBIDITY INDEX** |  |  |  |  |
| 0 | 107 (23.8) | 199 (31.4) | 1224 (41.0) | 1530 (37.6) |
| 1 | 165 (36.7) | 146 (23.0) | 606 (20.3) | 917 (22.5) |
| 2 | 74 (16.5) | 97 (15.3) | 572 (19.2) | 743 (18.3) |
| 3 | 103 (22.9) | 192 (30.3) | 584 (19.6) | 879 (21.6) |
| **TOBACCO HISTORY** |  |  |  |  |
| NEVER | < 10 | 18 (2.8) | 100 (3.3) | 124 (3.0) |
| PAST | 217 (48.3) | 285 (45.0) | 1303 (43.6) | 1805 (44.4) |
| CURRENT | 219 (48.8) | 311 (49.1) | 1522 (51.0) | 2052 (50.4) |
| UNKNOWN | < 10 | 20 (3.2) | 61 (2.0) | 88 (2.2) |
| **PRETREATMENT FEV1 (% PREDICTED)** |  |  |  |  |
| ≤30% | 51 (11.4) | 15 (2.4) | 15 (0.5) | 81 (2.0) |
| 31-50% | 136 (30.3) | 160 (25.2) | 229 (7.7) | 525 (12.9) |
| 51-80% | 200 (44.5) | 300 (47.3) | 1506 (50.4) | 2006 (49.3) |
| ≥80% | 62 (13.8) | 159 (25.1) | 1236 (41.4) | 1457 (35.8) |
| **HISTOLOGY** |  |  |  |  |
| ADENOCARCINOMA | 176 (39.2) | 369 (58.2) | 2699 (56.9) | 2244 (55.1) |
| SQUAMOUS CELL CARCINOMA | 202 (45.0) | 209 (33.0) | 964 (32.3) | 1375 (33.8) |
| UNKNOWN/OTHER | 71 (15.8) | 56 (8.8) | 323 (10.8) | 450 (11.1) |
| **GRADE** |  |  |  |  |
| 1 | 13 (2.9) | 80 (12.6) | 365 (12.2) | 458 (11.3) |
| 2 | 71 (15.8) | 313 (49.4) | 1457 (48.8) | 1841 (45.2) |
| 3 | 81 (18.0) | 168 (26.5) | 930 (31.1) | 1179 (29.0) |
| UNKNOWN | 284 (63.3) | 73 (11.5) | 234 (7.8) | 591 (14.5) |

| CLINICAL T CATEGORY |  |  |  |  |
| --- | --- | --- | --- | --- |
| T1A | 187 (41.6) | 395 (62.3) | 1329 (44.5) | 1911 (47.0) |
| T1B | 176 (39.2) | 168 (26.5) | 849 (28.4) | 1193 (29.3) |
| T2A | 86 (19.2) | 71 (11.2) | 808 (27.1) | 965 (23.7) |

**Table 3.** Descriptive statistics for head and neck cancer cohort. All variables listed in format of number of patients (column percentage). ECOG = Eastern Cooperative Oncology Group.

| COVARIATES | LOW INTENSITY TREATMENT<br>(N= 2951) | HIGH<br>INTENSITY TREATMENT (N= 4166) | OVERALL<br>(N= 7117) |
| --- | --- | --- | --- |
| **AGE** |  |  |  |
| <50 | 51 (1.7) | 170 (4.1) | 221 (3.1) |
| 50-59 | 652 (22.1) | 1378 (33.1) | 2030 (28.5) |
| 60-69 | 1299 (44.0) | 2134 (51.2) | 3433 (48.2) |
| 70-79 | 632 (21.4) | 425 (10.2) | 1057 (14.9) |
| ≥80 | 317 (10.7) | 59 (1.4) | 376 (5.3) |
| **BODY MASS INDEX** |  |  |  |
| UNDERWEIGHT (<18.5) | 715 (24.2) | 829 (19.9) | 1544 (21.7) |
| NORMAL (≥18.5, <25) | 1281 (43.4) | 1851 (44.4) | 3132 (44.0) |
| OVERWEIGHT (≥25, <30) | 566 (19.2) | 927 (22.3) | 1493 (21.0) |
| OBESE (≥30) | 260 (8.8) | 436 (10.5) | 696 (9.8) |
| UNKNOWN | 129 (4.4) | 123 (3) | 252 (3.5) |
| **RACE** |  |  |  |
| WHITE | 2350 (79.6) | 3429 (82.3) | 5779 (81.2) |
| BLACK | 524 (17.8) | 649 (15.6) | 1173 (16.5) |
| OTHER | 77 (2.6) | 88 (2.1) | 165 (2.3) |
| **SEX** |  |  |  |
| MALE | 2927 (99.2) | 4116 (98.8) | 7043 (99.0) |
| FEMALE | 24 (0.8) | 47 (1.1) | 71 (1.0) |
| OTHER | < 10 | < 10 | < 10 |
| **MARRIED** |  |  |  |
| NO | 1829 (8.8) | 2653 (63.7) | 4482 (63.0) |
| YES | 1122 (38.0) | 1513 (36.3) | 2635 (37.0) |
| **TOBACCO HISTORY** |  |  |  |
| NEVER | 259 (8.8) | 346 (8.3) | 605 (8.5) |
| PAST | 991 (33.6) | 1161 (27.9) | 2152 (30.2) |
| CURRENT | 1455 (49.3) | 2379 (57.1) | 3834 (53.9) |
| UNKNOWN | 246 (8.3) | 280 (6.7) | 526 (7.4) |
| **CHARLSON COMORBIDITY INDEX** |  |  |  |
| 0 | 1154 (39.1) | 2137 (51.3) | 3291 (46.2) |
| 1 | 594 (20.1) | 819 (19.7) | 1413 (19.9) |
| 2 | 499 (16.9) | 593 (14.2) | 1092 (15.3) |
| ≥3 | 704 (23.9) | 617 (14.8) | 1321 (18.6) |
| **PERFORMANCE STATUS (ECOG)** |  |  |  |
| 0 | 480 (16.3) | 928 (22.3) | 1408 (19.8) |
| 1 | 556 (18.8) | 877 (21.1) | 1433 (20.1) |
| 2 | 146 (4.9) | 128 (3.1) | 274 (3.8) |
| 3 | 54 (1.8) | 17 (0.4) | 71 (1.0) |
| 4 | < 10 | < 10 | 10 (0.1) |
| UNKNOWN | 1706 (57.8) | 2215 (53.2) | 3921 (55.1) |
| **PRIMARY SITE** |  |  |  |
| HYPOPHARYNX | 309 (10.5) | 396 (9.5) | 705 (9.9) |
| LARYNX | 794 (26.9) | 1057 (25.4) | 1851 (26.0) |
| ORAL CAVITY | 178 (6.0) | 129 (3.1) | 307 (4.3) |
| OROPHARYNX | 1670 (56.6) | 2584 (62.0) | 4254 (59.8) |
| **P16 STATUS** |  |  |  |
| POSITIVE | 259 (8.8) | 421 (10.1) | 680 (9.6) |
| NEGATIVE | 144 (4.9) | 231 (5.5) | 375 (5.3) |
| NOT TESTED | 2548 (86.3) | 3514 (84.3) | 6062 (85.2) |
| **T CATEGORY** |  |  |  |
| 1 | 345 (11.7) | 493 (11.8) | 838 (11.8) |
| 2 | 879 (29.8) | 1296 (31.1) | 2175 (30.6) |
| 3 | 975 (33.0) | 1317 (31.6) | 2292 (32.2) |
| 4 | 752 (25.5) | 1060 (25.4) | 1812 (25.5) |
| **N CATEGORY** |  |  |  |
| 0 | 556 (18.8) | 572 (13.7) | 1128 (15.8) |
| 1 | 568 (19.2) | 617 (14.8) | 1185 (16.7) |
| 2 | 1684 (57.1) | 2796 (67.1) | 4480 (62.9) |
| 3 | 143 (4.8) | 181 (4.3) | 324 (4.6) |

**Figure. 1.**
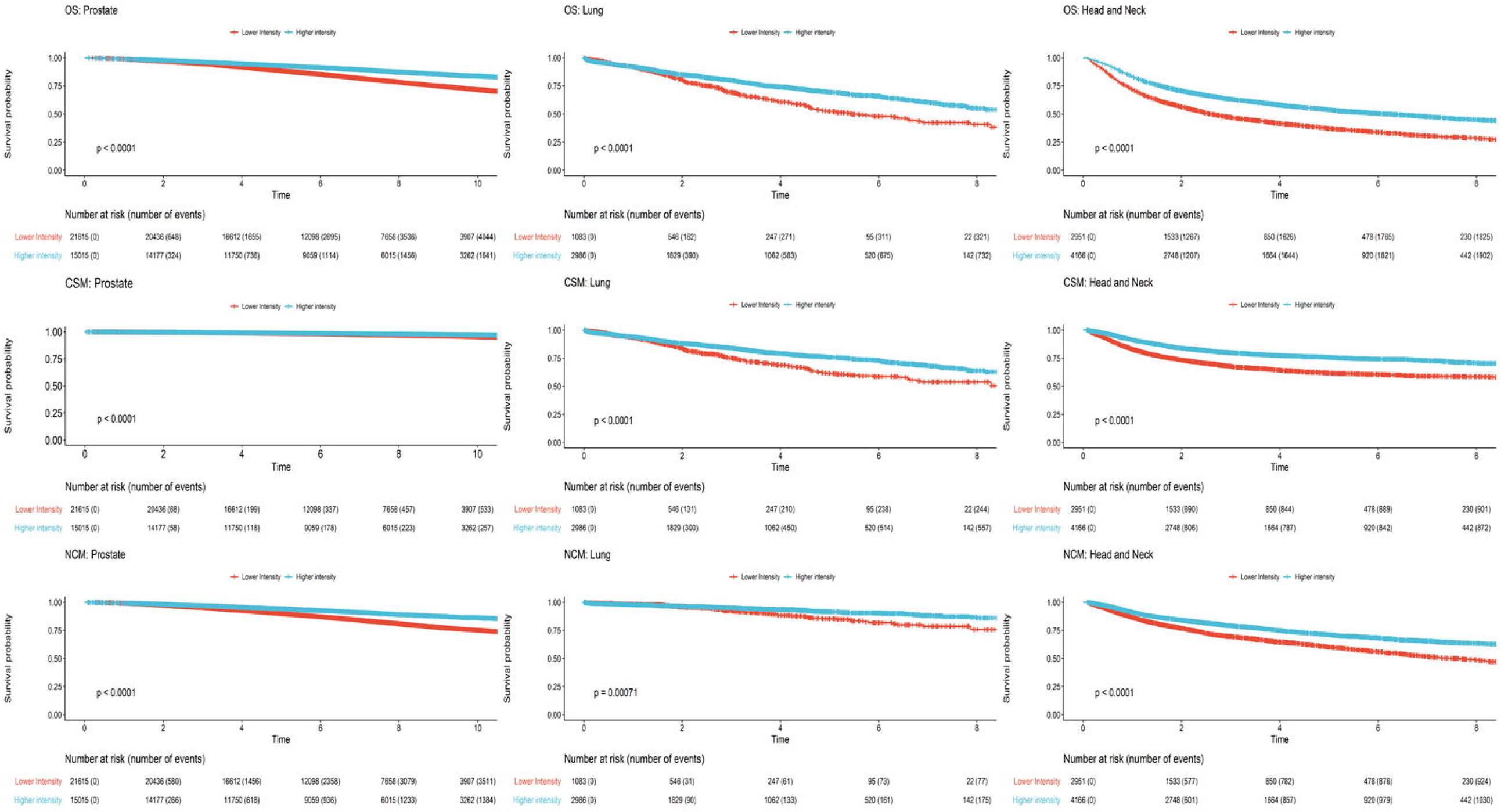
Kaplan-Meier survival curves showing unadjusted associations between treatment strategies for the prostate cancer (top), lung cancer (middle), and head and neck cancer (bottom) cohorts. OS: overall survival. CSM: cancer-specific mortality. NCM: non-cancer mortality. Prostate cancer cohort compares radical prostatectomy (blue) vs. definitive radiotherapy (red). Lung cancer cohort compares lobectomy (blue) vs. sub-lobar resection or definitive radiotherapy (red). Head and neck cancer cohort compares definitive radiotherapy with induction and/or concurrent cisplatin-based chemotherapy (blue) vs. definitive radiotherapy alone or with alternative systemic therapy (red).

For prostate cancer patients, prostatectomy was associated with significantly reduced CSM compared to RT after adjusting for covariates using IPTW (hazard ratio (HR) 0.82, 95% confidence interval (CI): 0.64, 0.92; p=0.02) (**Figure 1; Table 4**). Prostatectomy was also associated with significantly reduced *non-cancer mortality* (HR 0.71, 95% CI: 0.65, 0.74; (p<0.001) and improved OS (HR 0.73, 95% CI: 0.66, 0.75; p<0.001) with IPTW (**Figure 1**; **Table 4**). After correction using BRACE, however, the effect on OS was attenuated substantially (HR 0.98; 95% CI: 0.95, 1.00) with the upper bound of the 95% CI including the null (**Table 4**). For additional validation, we analyzed a similar cohort of early stage prostate cancer patients from a recently published analysis of 50,804 low-risk prostate cancer patients from SEER data (courtesy, Dr. Benjamin Kann and Dr. Joseph Miccio);^15^ the adjusted effect of prostatectomy relative to RT before (with IPTW) and after BRACE correction were HR 0.54 (95% CI: 0.50, 0.61) and 0.97 (95% CI: 0.95, 0.99), respectively.

**Table 4.** Effects of intensive treatment approaches on cancer-specific mortality, non-cancer mortality, and overall survival for each clinical cohort. Results are presented from unadjusted, Cox multivariable (MVA), and inverse probability treatment weighting (IPTW) models. The Bias Reduction through Analysis of Competing Events (BRACE) correction was applied to IPTW estimates. *statistically significant with p < 0.05, **statistically significant with p < 0.001. †Note that the BRACE method does not result in a p value. ⱡReference values were obtained from randomized trials and meta-analyses (^a^surgery vs. active monitoring; ^b^radiotherapy vs. active monitoring; ^c^lobectomy vs. segmentectomy; ^d^locoregional therapy with chemotherapy vs. locoregional therapy alone).^32-34^

| Outcome | Prostate Cancer | Lung Cancer | Head and Neck Cancer |
| --- | --- | --- | --- |
| **Cancer-Specific Mortality** |  |  |  |
| Unadjusted | 0.65 (0.57, 0.75)** | 0.66 (0.56, 0.76)** | 0.58 (0.53, 0.63)** |
| MVA | 0.85 (0.73, 0.99)* | 0.78 (0.66, 0.92)* | 0.65 (0.59, 0.72)** |
| IPTW | 0.82 (0.64, 0.92)* | 0.76 (0.61, 0.88)* | 0.64 (0.58, 0.71)** |
| **Non-Cancer Mortality** |  |  |  |
| Unadjusted | 0.54 (0.51, 0.57)** | 0.63 (0.48, 0.83)** | 0.64 (0.59, 0.70)** |
| MVA | 0.73 (0.68, 0.78)** | 0.83 (0.62, 1.11) | 0.76 (0.69, 0.84)** |
| IPTW | 0.71 (0.65, 0.74)** | 0.87 (0.64, 1.17) | 0.76 (0.69, 0.84)** |
| **Overall Survival** |  |  |  |
| Unadjusted | 0.55 (0.52, 0.59)** | 0.65 (0.57, 0.74)** | 0.61 (0.57, 0.65)** |
| MVA | 0.75 (0.71, 0.80)** | 0.79 (0.68, 0.91)* | 0.71 (0.66, 0.76)** |
| IPTW | 0.73 (0.66, 0.75)** | 0.79 (0.66, 0.89)* | 0.70 (0.66, 0.76)** |
| BRACE | 0.98 (0.95, 1.00)† | 0.81 (0.65, 0.94)† | 0.81 (0.76, 0.86)† |
| **Reference Values<sup>‡</sup>** |  |  |  |
|  | 0.93 (0.65, 1.35)<sup>a</sup> | 0.72 (0.48, 1.09)<sup>c</sup> | 0.88 (0.85, 0.92)<sup>d</sup> |
|  | 0.94 (0.65, 1.36)<sup>b</sup> |  |  |

For the lung cancer cohort, lobectomy was similarly associated with significantly improved CSM (HR 0.76, 95% CI: 0.61, 0.88; p=0.02), NCM (HR 0.87, 95% CI 0.64, 1.17; p=0.43), and OS (HR 0.79, 95% CI: 0.66, 0.89; p<0.01), after adjusting for covariates (**Figure 1; Table 4**). After BRACE application, the estimated effect on survival was slightly attenuated (HR 0.81, 95% CI: 0.65, 0.94). For the head/neck cancer cohort, intensive chemotherapy was also associated with significantly improved CSM (HR 0.64, 95% CI: 0.58, 0.71; p<0.001), NCM (HR 0.76, 95% CI: 0.69, 0.84; p<0.001), and OS (HR 0.70, 95% CI: 0.66, 0.76; p < 0.001), with an attenuated effect on OS with BRACE (HR 0.81, 95% CI: 0.76, 0.86) (**Figure 1**; **Table 4**).

When the proportional hazards assumption was not met, we modeled treatment as a time-varying covariate. **Supplemental Table 1** shows results for time periods over which the assumption held. In general, the corrected OS estimates were nearly identical, but in the prostate cancer cohort, the null hypothesis was not rejected using BRACE, and in the lung cancer cohort, the standard and BRACE estimates diverged more, indicating sensitivity to the proportional hazards assumption. Using Monte Carlo methods for confidence interval estimation, the BRACE-adjusted effects of intensive treatment on OS for the prostate, head/neck, and lung cancer cohorts were HR 0.98 (95% CI: 0.90, 1.07), HR 0.81 (95% CI: 0.74, 0.91), and HR 0.81 (95% CI 0.76, 0.89), respectively.

## DISCUSSION

Typical strategies to address treatment selection bias in observational data include multivariable Cox proportional hazards regression and propensity score modeling.^26-30^ While valuable, these methods may be insufficient to eliminate residual confounding, leading to erroneous inferences.^1-8^ Competing risks analysis can diagnose residual confounding by identifying mechanistically implausible effects of treatment on competing health events.^11,12^ In each cohort we examined, while there were strong associations between intensive treatment and improved survival, competing risks analysis revealed these effects were driven in part by associations with reduced non-cancer mortality, even after adjusting for numerous measurable confounders. The likely explanation for this is the selective use of intensive treatment in patients with more favorable baseline health characteristics, thus leading to reduced non-cancer mortality, rather than effects on the competing event *per se*. However, simply identifying this problem does not inherently provide a method to address it.

Here we applied a recently described method to attenuate bias, which was previously shown to result in lower model error compared to standard approaches in simulated data.^13^ While true effects are not directly observable in non-randomized studies, multiple applications of BRACE to clinical cohorts yielded attenuated treatment effect estimates more consistent with high-level evidence than uncorrected estimates. Such comparisons should be viewed with caution, given methodological and population differences across studies, but they can lend insight when comparing potentially biased results.

For example, the randomized ProtecT trial found no difference in OS by treatment in 1,643 patients with predominantly low-risk prostate cancer,^31^ which our results generally support. While ProtecT did not directly quantify the effect of prostatectomy vs. radiotherapy, both were compared to a common control group (active monitoring), with nearly identical effects (**Table 4**). Similarly, the MACH-NC meta-analysis, which investigated the effect of chemotherapy in addition to RT for 16485 patients across 87 randomized trials, reported an effect of chemotherapy on survival (HR 0.88), close to our BRACE-adjusted estimate.^32^ While evidence regarding the comparative effectiveness of treatments for early stage NSCLC are conflicting,^33-41^ our results indicated a survival advantage to lobectomy after BRACE correction. Though large randomized trials are lacking, in the meta-analysis by Zheng et al., the effect was attenuated in trials with higher levels of evidence.^33^ Of note, in some studies, lobectomy has been associated with higher 90-day mortality compared to stereotactic ablative radiotherapy (SABR);^41^ if true, BRACE would under-correct for treatment selection bias.

Our findings thus have important implications regarding analyses using observational data. For example, the National Cancer Database lacks cause-specific event data, precluding the application of BRACE and leaving many analyses vulnerable to undiagnosed bias. Missing or inadequate comorbidity data may also contribute to residual confounding; application of BRACE in databases that include cause-specific event data could help mitigate this problem proactively.

This study has several limitations. Notably, the proportional relative hazards model treats several key quantities as independent, a strong condition that is not always verifiable. In our analysis of clinical data, it is important to note the corrected estimates could still be biased. Furthermore, inferences can be sensitive to the proportional hazards assumption or to the method used to estimate confidence intervals, especially when close to the null. Moreover, gains using BRACE depend on leveraging a critical assumption: namely, that treatment does not reduce the hazard for non-cancer events. While this is generally valid when comparing more vs. less intensive treatments (e.g., A vs. A+B designs), in other contexts it may not be possible to bound the effects of a treatment on competing events, such as when comparing two systemic therapies).

In summary, we present the clinical application of a novel method (BRACE) to mitigate bias from residual confounding. Appropriate application in observational, non-randomized data would likely improve effect estimation and inferences.

## Data Availability

Data will be made available upon request.

## Data Sharing and Availability

VINCI and SEER data supporting the findings of this study are available from the U.S. Veterans Affairs Administration and SEER, respectively, but restrictions apply to the availability of these data, which were used under license for the current study, and so are not publicly available. Data are however available from the authors upon reasonable request and with permission of these authorities.

## ACKNOWLEDGEMENTS

The authors wish to thank James Murphy for facilitating access to VINCI data, and Joseph Miccio and Benjamin Kann for facilitating access to SEER data.

---

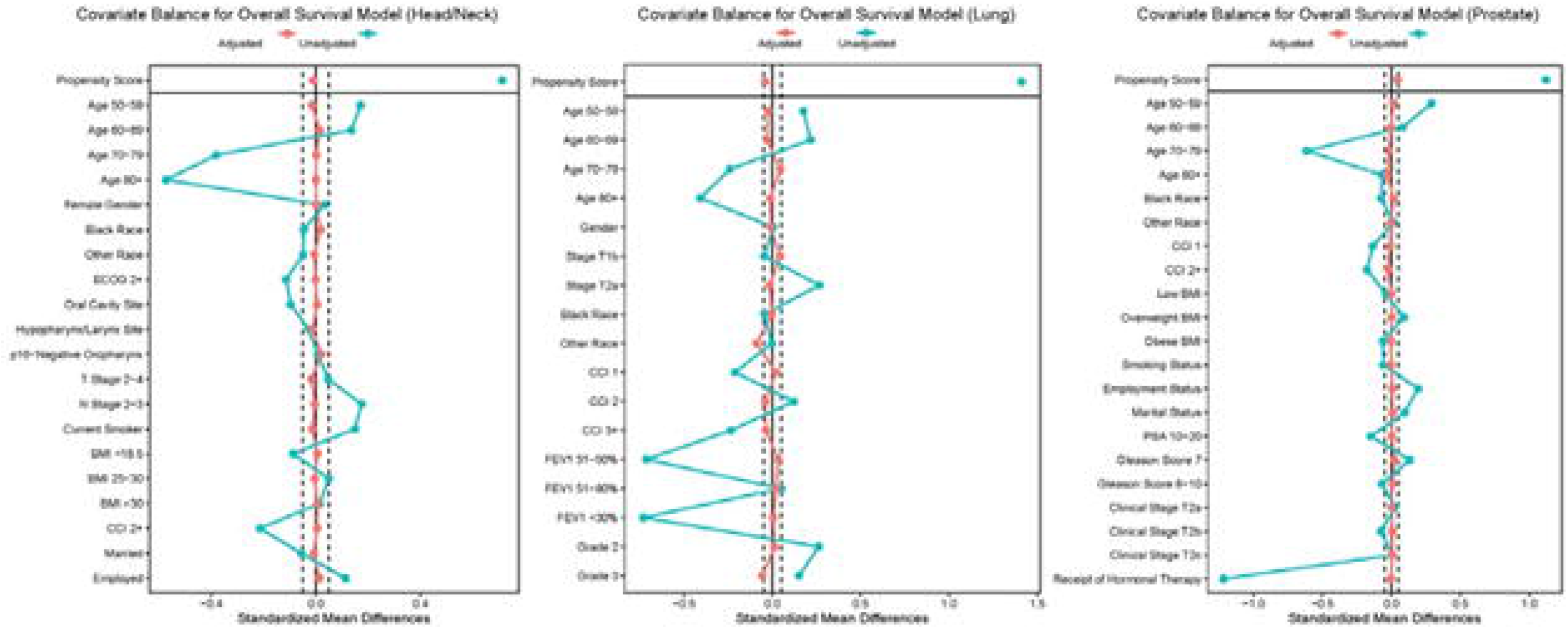

## SUPPLEMENTARY DATA

**Supplementary Table 1.**
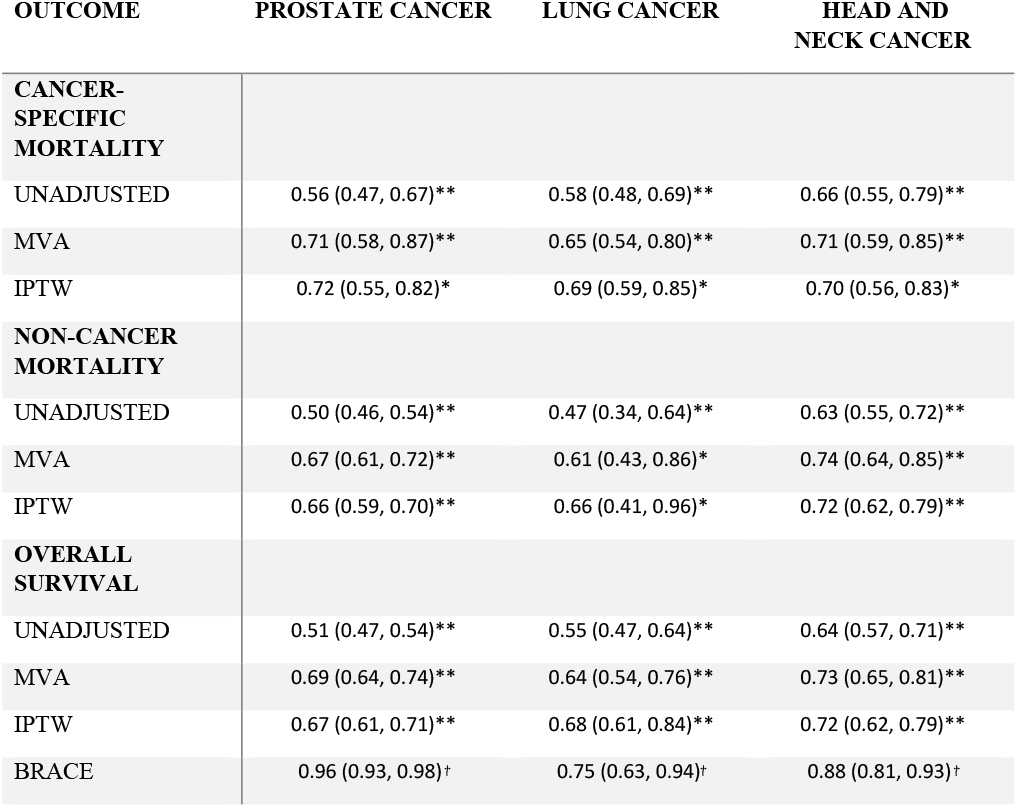
Effects of intensive treatment approaches for time periods during which the proportional hazards assumption was found to be valid (for prostate cancer, 3.1-10.4 years, with the time point of 10.4 years chosen based on published literature25; for lung cancer, excluding the first year of follow-up; for head and neck cancer, 2-10 years). Results are presented from unadjusted, multivariable Cox (MVA), and inverse probability treatment weighting (IPTW) models. The Bias Reduction through Analysis of Competing Events (BRACE) correction was applied to IPTW estimates. *statistically significant with p < 0.05, **statistically significant with p < 0.001. †Note that the BRACE method does not result in a p value.

## REFERENCES

1. Becher, H. The concept of residual confounding in regression models and some applications. Stat. Med. 11, 1747–58 (1992).

2. Jatoi, I. Residual confounding threatens the validity of observational studies on breast cancer local therapy. Cancer 126, 2317–2020 (2020).

3. Harrell F. E. Jr., Lee K. L. & Mark D. B. Multivariable prognostic models: issues in developing models, evaluating assumptions and adequacy, and measuring and reducing errors. Stat. Med. 15, 361–387 (1996).

4. Bosco J. L., et al. A most stubborn bias: no adjustment method fully resolves confounding by indication in observational studies. J. Clin. Epidemiol. 63, 64–74 (2010).

5. Giordano S. H., et al. Limits of observational data in determining outcomes from cancer therapy. Cancer 112, 2456–66 (2008).

6. McGale P., et al. Can observational data replace randomized trials? J. Clin. Oncol. 34, 3355–3357 (2016).

7. Kumar A., et al. Evaluation of the use of cancer registry data for comparative effectiveness research. JAMA Netw. Open 3, e2011985 (2020).

8. Soni P. D., et al. Comparison of population-based observational studies with randomized trials in oncology. J. Clin. Oncol. 37, 1209–1216 (2019).

9. Korn E. L. & Freidlin B. Methodology for comparative effectiveness research: potential and limitations. J. Clin. Oncol. 30, 4185–4187 (2012).

10. Hahn O. M. & Schilsky R. L. Randomized controlled trials and comparative effectiveness research. J. Clin. Oncol. 30, 4194–4201 (2012).

11. Mell L. K. & Jeong J. H. Pitfalls of using composite primary end points in the presence of competing risks. J. Clin. Oncol. 28, 4297–4299 (2010).

12. Mell L. K., et al. Cause-specific effects of radiotherapy and lymphadenectomy in stage I-II endometrial cancer: a population-based study. J. Natl. Cancer Inst. 105, 1656–1666 (2013).

13. Mell LK, Nelson TJ, Thompson CA, Williamson CW, Vitzthum LK, Zou J. Bias Reduction through Analysis of Competing Events (BRACE): A Novel Method to Mitigate Bias from Residual Confounding in Observational Data. medRxiv. 2020 Jan 1.

14. American College of Surgeons. Facility Oncology Registry Data Standards. https://www.facs.org/-/media/files/quality-programs/cancer/ncdb/fords-2016.ashx (2016).

15. Miccio, J. A. et al. Quantifying treatment selection bias effect on survival in comparative effectiveness research: findings from low-risk prostate cancer patients. Prostate Cancer Prostatic Dis. In press (2020) doi:10.1038/s41391-020-00291-3.

16. Bryant, A. K. et al. Stereotactic body radiation therapy versus surgery for early lung cancer among US veterans. Ann. Thorac. Surg. 105, 425–431 (2018).

17. Templ M, Kowarik A, Filzmoser P. Iterative stepwise regression imputation using standard and robust methods. Comput Stat Data Anal. 2011 Oct 1;55(10):2793–806.

18. Vitzthum, L. K. et al. Selection of head and neck cancer patients for intensive therapy. Int. J. Radiat. Oncol. Biol. Phys. 106, 157–166 (2020).

19. Stein, A. P. et al. Prevalence of human papillomavirus in oropharyngeal squamous cell carcinoma in the United States across time. Chemical Research in Toxicology 27, 462–9 (2014).

20. Fakhry, C. et al. The prognostic role of sex, race, and human papillomavirus in oropharyngeal and nonoropharyngeal head and neck squamous cell cancer. Cancer 123, 1566–1575 (2017).

21. Carmona, R. et al. Validated competing event model for the stage I-II endometrial cancer population. Int. J. Radiat. Oncol. Biol. Phys. doi:10.1016/j.ijrobp.2014.03.047 (2014).

22. Zakeri, K., Rose, B. S., Gulaya, S., D’Amico, A. V. & Mell, L. K. Competing event risk stratification may improve the design and efficiency of clinical trials: Secondary analysis of SWOG 8794. Contemp. Clin. Trials doi:10.1016/j.cct.2012.09.008.(2013)

23. Carmona, R. et al. Improved method to stratify elderly patients with cancer at risk for competing events. J. Clin. Oncol. doi:10.1200/JCO.2015.65.0739 (2016).

24. Mell, L. K. et al. Nomogram to Predict the Benefit of Intensive Treatment for Locoregionally Advanced Head and Neck Cancer. Clin. Cancer Res. doi:10.1158/1078-0432.CCR-19-1832. (2019)

25. Mell, L. K. et al. Effect of hormone therapy within risk groups defined by generalized competing event model: ancillary analysis of NRG Oncology’s RTOG 9408 (abstr.) Int. J. Radiat. Oncol. Biol. Phys. 108, E863–E864 (2020) doi:10.1016/j.ijrobp.2020.07.435

26. Therneau T. M. & Grambsch P. M. The Cox Model. in Modeling Survival Data: Extending the Cox Model. Statistics for Biology and Health (eds. Therneau T.M. & Grambsch P. M.) 39–77 (Springer, 2000).

27. Rosenbaum, P. R. & Rubin, D. B. The central role of the propensity score in observational studies for causal effects. Biometrika 70, 41–55 (1983).

28. Austin, P. C. An introduction to propensity score methods for reducing the effects of confounding in observational studies. Multivariate Behav. Res. 46, 399–424 (2011).

29. Austin, P. C. A tutorial and case study in propensity score analysis: An application to estimating the effect of in-hospital smoking cessation counseling on mortality. Multivariate Behav. Res. 46, 119–151 (2011).

30. Austin, P. C. The performance of different propensity-score methods for estimating relative risks. J. Clin. Epidemiol. 61, 537–545 (2008).

31. Hamdy, F. C. et al. 10-year outcomes after monitoring, surgery, or radiotherapy for localized prostate cancer. N. Engl. J. Med. 375, 1415–24 (2016).

32. Pignon, J. P., le Maître, A., Maillard, E. & Bourhis, J. Meta-analysis of chemotherapy in head and neck cancer (MACH-NC): An update on 93 randomised trials and 17,346 patients. Radiother. Oncol. 92, 4–14 (2009).

33. Zheng, Y. Z. et al. Oncologic outcomes of lobectomy vs. segmentectomy in non-small cell lung cancer with clinical T1N0M0 stage: a literature review and meta-analysis. J. Thorac. Dis. 12, 3178–3187 (2020).

34. Razi, S. S., John, M. M., Sainathan, S. & Stavropoulos, C. Sublobar resection is equivalent to lobectomy for T1a non–small cell lung cancer in the elderly: a Surveillance, Epidemiology, and End Results database analysis. J. Surg. Res. 200, 683–689 (2016).

35. Ginsberg, R. J. & Rubinstein, L. V. Randomized trial of lobectomy versus limited resection for T1 N0 non-small cell lung cancer. Ann. Thorac. Surg. 60, 615–22 (1995).

36. Khullar, O. V. et al. Survival after sublobar resection versus lobectomy for clinical stage IA lung cancer: an analysis from the National Cancer Data Base. J. Thorac. Oncol. 10, 1625–33 (2015).

37. El-Sherif, A. et al. Outcomes of sublobar resection versus lobectomy for stage I non-small cell lung cancer: a 13-year analysis. Ann. Thorac. Surg. 82, 408–15 (2006).

38. Chang, J. Y. et al. Stereotactic ablative radiotherapy versus lobectomy for operable stage I non-small-cell lung cancer: A pooled analysis of two randomised trials. Lancet Oncol. 16, 630–7 (2015).

39. Zhang, B. et al. Matched-pair comparisons of stereotactic body radiotherapy (SBRT) versus surgery for the treatment of early stage non-small cell lung cancer: A systematic review and meta-analysis. Radiother. Oncol. 112, 250–5 (2014).

40. Zheng, X. et al. Survival outcome after stereotactic body radiation therapy and surgery for stage I non-small cell lung cancer: a meta-analysis. Int. J. Radiat. Oncol. Biol. Phys. 90, 603–11 (2014).

41. Shirvani S. M., et al. Lobectomy, sublobar resection, and stereotactic ablative radiotherapy for early-stage non-small cell lung cancers in the elderly. JAMA Surg. 149, 1244–53 (2014).

